# The Role of Air Conditioning in the Diffusion of Sars-CoV-2 in Indoor Environments: a First Computational Fluid Dynamic Model, based on Investigations performed at the Vatican State Children’s Hospital

**DOI:** 10.1101/2020.08.25.20181420

**Authors:** Luca Borro, Lorenzo Mazzei, Massimiliano Raponi, Prisco Piscitelli, Alessandro Miani, Aurelio Secinaro

## Abstract

**Background:** About 15 million people worldwide were affected by the Sars-Cov-2 infection, which already caused 600,000 deaths. This virus is mainly transmitted through exhalations from the airways of infected persons, so that Heating, Ventilation and Air Conditioning (HVAC) systems might play a role in increasing or reducing the spreading of the infection in indoor environments.

**Methods:** We modelled the role of HVAC systems in the diffusion of the contagion through Computational Fluid Dynamics (CFD) simulations of cough at the “Bambino Gesù” Vatican State Children’s Hospital.

Both waiting and hospital rooms were modeled as indoor scenarios. A specific Infection-Index (η) parameter was used to estimate the amount of contaminated air inhaled by each person present in the simulated indoor scenarios. The potential role of exhaust air ventilation systems placed above the coughing patient’s mouth was also assessed.

**Results:** Our CFD-based simulations of the waiting room show that HVAC air-flow remarkably enhances infected droplets diffusion in the whole indoor environment within 25 seconds from the cough event, despite the observed dilution of saliva particles containing the virus. At the same time also their number is reduced due to removal through the HVAC system or deposition on the surfaces. The proper use of Local Exhaust Ventilation systems (LEV) simulated in the hospital room was associated to a complete reduction of infected droplets spreading from the patient’s mouth in the first 0.5 seconds following the cough event. In the hospital room, the use of LEV system completely reduced the η index computed for the patient hospitalized at the bed next to the spreader, with a decreased possibility of contagion.

**Conclusions:** CFD-based simulations for indoor environment can be useful to optimize air conditioning flow and to predict the contagion risk both in hospitals/ambulatories and in other public/private settings.

## Introduction

In early 2020 the world was affected by a significant pandemic caused by infection with Sars-Cov-2, a Coronavirus responsible for the Covid-19 disease, which causes a severe respiratory syndrome that can lead to death in the absence of an effective drug and vaccine treatment. Coronaviruses are single-strained positive-sense RNA viruses that include seasonal common cold viruses (229E, OC43, NL63, HKU-1), but they also include more virulent viruses like as Mers-Cov and Sars-Cov. The Sars-Cov coronaviruses are responsible for severe acute respiratory syndrome, while the Mers-Cov are responsible for the Middle-East Respiratory Syndrome. The Coronavirus pandemic caused over 15 million infections, with more than 619,000 deaths worldwide. Asia with China, Europe, USA, South-America and Mexico are the continents most affected by the pandemic. The Sars-Cov-2 can spread through the air over large distances [1] and viral particles have also been found on particulate matter [2]. The airways are, therefore, the primary way of transmission through the droplets and aerosol dispersion in the environment [3]. A less critical way of transmission is direct contact with hands and contaminated objects [4]. To make clear the scientific treatment of the contagion transmission in epidemic contexts, some relevant articles about the nomenclature have been published over the years concerning the definition of droplets and droplets nuclei. All these particles can be expelled from the mouth or the nose through sneezing, coughing and talking. Droplet transmission is a way of contagion consisting in expelled particles from the mouth that quickly settle to the ground under the gravity force. The droplets nuclei transmission is the transmission that occurs by expelled particles smaller in size compared to the droplets. The World Health Organization has established that particles with diameters less than 5 μm are called droplets nuclei, while particles with diameters greater than 5 μm are called droplets [5]. The large particles typically settle within 1 meter of the source but this value depends on several external dynamic factors that make it susceptible to changes. The smaller particles instead remain suspended in the air for varying periods and evaporate much more quickly.

Nicas and Jones 2010 [6] have proposed a useful classification dividing the particles into “Respirable Particles” and “Inspirable Particles”. The respirable particles have a diameter of 10 μm or less and deposit throughout the respiratory tract, including the alveolar region, whereas inspirable particles with a diameter between 10 μm and 100 μm deposit in the upper airways and tracheobronchial regions only [7].

Several studies have analyzed the spread of viral infections caused by inhaling viral particles contained in airborne biological material as droplets or droplets nuclei. The dispersion of droplets and droplets nuclei (aerosols) has considerable relevance in the spread of various types of viruses like as Influenza A that are often present in small particles (< 5 μm), Sars-Cov and Sars-Cov-2 [9]. In 2018 Yang et al. carried out in-depth studies on the size of the droplets and nuclei droplets during coughing. In this study [10] 82% of the nuclei droplets belonged to the range of diameters between 0.74 μm to 2.12 μm while the coughed droplets showed three peaks of diameters approximately at 1, 2 and 8 μm.

All previous experimental studies on this topic give us much complete information about the physical-mechanical characterization of cough and free breathing. The dispersion of droplets and aerosols indoors is the main responsible for respiratory viral infections and its control is crucial for public health, especially in public settings such as hospitals, offices and recreational places.

The purpose of this work is to analyze the influence of Heating, Ventilation and Air Conditioning (HVAC) systems on the air-dispersion of aerosols generated by a cough within a closed environment.

## Materials and Methods

The simulation consists in two differents cases: Case 1 and Case 2. Case 1 consists of a waiting room with a surface of 65 m^2^, in which the air is assumed to be at a temperature of 26 °C and a relative humidity of 50%. The ground, ceiling are lateral walls are treated as adiabatic.

For Case 1 (Figure 1), we produced three different scenarios, considering no HVAC (Scenario A) and an active HVAC system with two different levels of volume flow rate: nominal flow rate (Scenario B) and double flow rate (Scenario C). The simulation involving a two-fold increase in volume flow rate is aimed at verifying its effect on the spreading of contaminated air and droplets within the room. The general assumption is that this effect reduces their concentration of droplets by ensuring a greater air recirculation in less time than the HVAC system at a nominal flow rate [11]. The objective of this analysis is to investigate the role played by the HVAC system and its intensity on the spreading.

**Figure 1:**
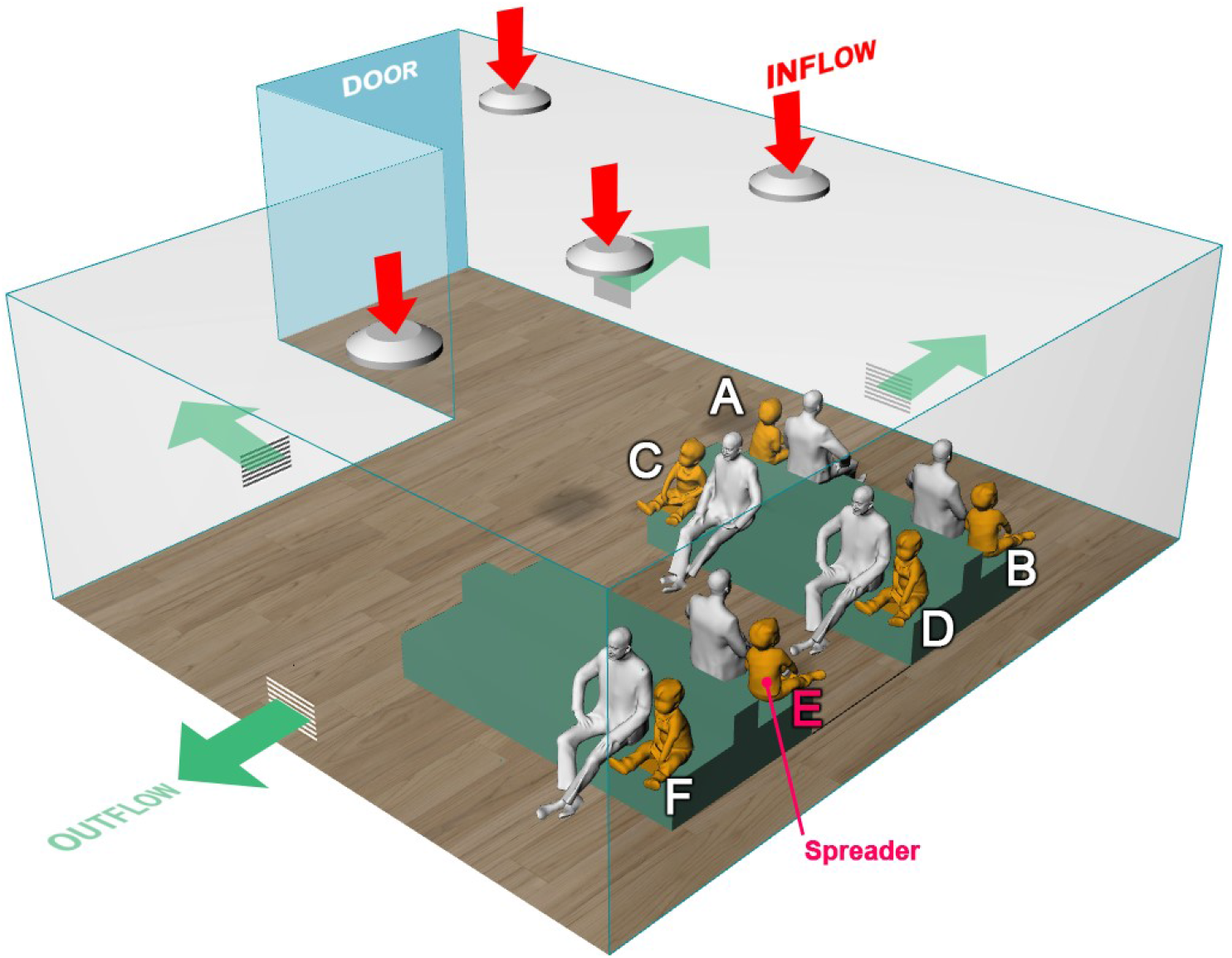
Case 1, a pediatric emergency waiting room with 6 pairs of men and children. The spreader is represented by kid E.

The room includes 4 HVAC inflows and 4 HVAC outflows. The HVAC round diffusers have been 3D modelled to ensure a physically correct air direction in the room. The diffusers have a diameter of 77 cm with two concentric cones with 46 cm and 20 cm respectively. The HVAC return grilles were installed on the lower part of the lateral walls at 25 cm from the floor. For the HVAC system operating at nominal conditions, 2020 m^3^/h enter from each diffuser, while the return grilles ensure a recirculation of 1790 m^3^/h each. For Scenario C, the two-fold increase results in an air flow of 4040 m^3^/h from each inlet and 3580 m^3^/h extracted by each return grill.

In all the scenarios of Case 1 the simulations include six adult men and six young childrens sit on benches modelled with manikins obtained by 3D online library [12] [13], with a body temperature of 31 °C [14]. The six pairs of men and children were tagged with a letter from A to F, where the child E represents the spreader subject that triggers the cough event. All people sit opposite each other and the distance from one person to another is about 1.70 m.

The Case 2 (Figure 2) consists of a children’s hospital room with two hospitalized patients at body temperature of 31 °C [14]. The room is assumed to be at a temperature of 26 °C and a relative humidity of 50%. The ground, ceiling and lateral walls are adiabatic. The distance between the two patients is 1.90 m.

**Figure 2:**
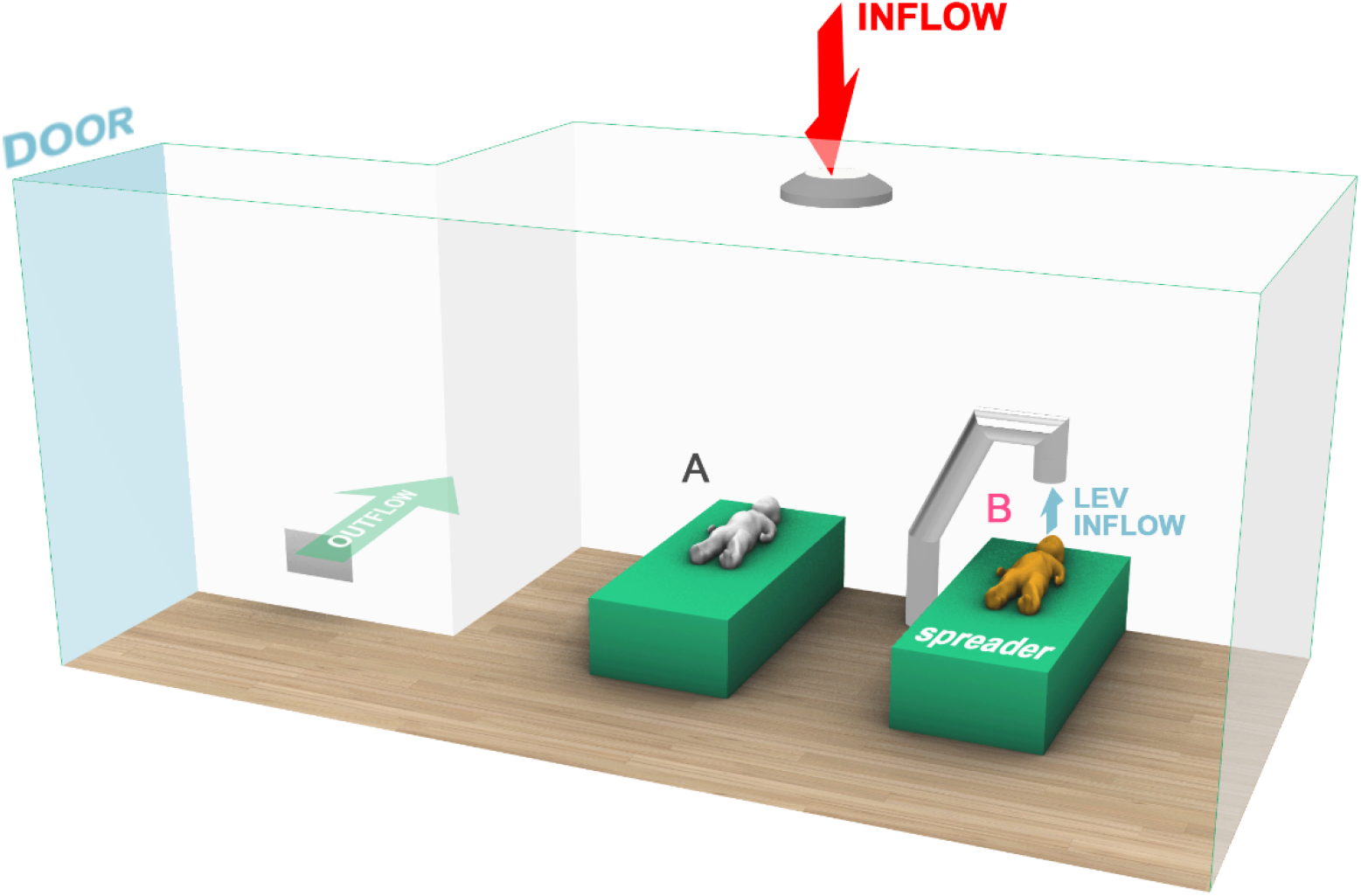
Case 2, an hospital recovery room with two beds and two pediatric patients. The LEV system acts directly above the spreader subject (Kid B).

For Case 2, two different scenarios were considered. Scenario A represents the baseline situation, in which the subject B is the spreader, whereas contaminated air and droplets are diffused and convected due to buoyancy, gravity and the HVAC system. In the additional Scenario B, a Local Exhaust Ventilation (LEV) system was placed above the spreader’s face. It consists of a tube connected to an air treatment unit equipped with different types of high-grade air filters. The LEV system is commonly employed in the industry to ensure and improve the air quality of workspaces. The objective of this analysis is to analyze the change in spreading due to the presence of the local exhaust system.

In our scenario a simplified LEV system was modeled considering a circular section tube with a diameter of 20 cm and an inlet area of 300 cm^2^. The Local Exhaust system has an inlet air flow of 1080 m^3^/h [15] and is positioned on the bed at about 43 cm above the spreader patient’s mouth (Figure 2). Additionally, an HVAC system is present in the room, with an inlet air flow of 480 m^3^/h and an outflow through the return grille of 420 m^3^/h [16].

Considering the CFD setup, the simulations were carried out with the commercial CFD solver ANSYS Fluent release 2019 R3. The medium was modeled as an incompressible ideal gas with constant properties calculated at ambient conditions. The gravity was included to account for the effects of buoyancy due to the temperature difference among ambient, manikins superficial temperature and sneezed air.

As far as the coughing process goes, it is worth underlining how appropriate modeling of the droplet transport plays a key role in determining the consequences of the coughing event. For this purpose, a literature review was carried out to determine the most reliable setup for the scope of work, in terms of both modeling approach and boundary conditions.

Coupled Eulerian-Lagrangian simulations represent for at least 15 years the state-of-the-art for the computational study of the aerial dispersion of contaminants [17]–[23]. An unsteady particle tracking was used, to include the effect of secondary break-up through a TAB model. The evaporation was also accounted for, considering a volatile component fraction of 94% [24]. The effects of the turbulent flow on the droplets are with Discrete Random Walk approach.

A poly-dispersed droplet population was considered to make the most of the capabilities of the Lagrangian approach. Among the many possible size distribution reported in the literature for a coughing event, it was decided to exploit the setup described by Bi et al., 2018 [22], who used the size distribution from Duguid, 1946 (see Table 1). Such a population was modeled by fitting the data with a Rosin-Rammler distribution.

**Table 1:**
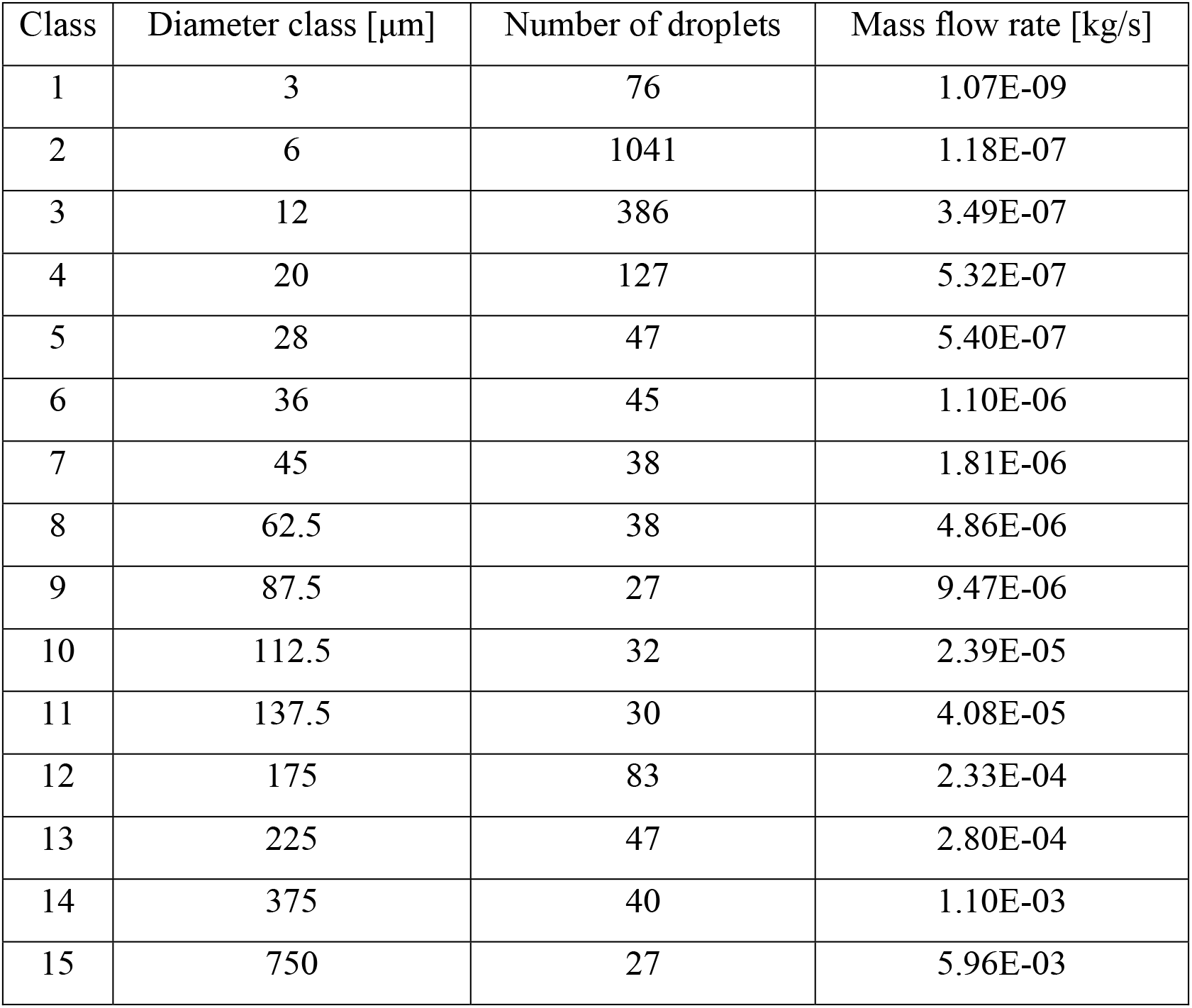
Droplet size distribution from Duguid (1946)

| Class | Diameter class [$\mu\text{m}$] | Number of droplets | Mass flow rate [$\text{kg/s}$] |
| --- | --- | --- | --- |
| 1 | 3 | 76 | 1.07E-09 |
| 2 | 6 | 1041 | 1.18E-07 |
| 3 | 12 | 386 | 3.49E-07 |
| 4 | 20 | 127 | 5.32E-07 |
| 5 | 28 | 47 | 5.40E-07 |
| 6 | 36 | 45 | 1.10E-06 |
| 7 | 45 | 38 | 1.81E-06 |
| 8 | 62.5 | 38 | 4.86E-06 |
| 9 | 87.5 | 27 | 9.47E-06 |
| 10 | 112.5 | 32 | 2.39E-05 |
| 11 | 137.5 | 30 | 4.08E-05 |
| 12 | 175 | 83 | 2.33E-04 |
| 13 | 225 | 47 | 2.80E-04 |
| 14 | 375 | 40 | 1.10E-03 |
| 15 | 750 | 27 | 5.96E-03 |

The population is injected in the time range of 0.042-0.136 s, which represents only a fraction of the temporal velocity profile applied to the eulerian phase at the mouth of the spreader subject via a velocity-inlet condition (Figure 3). The droplets have velocity and temperature of the consistent with the contaminated air injected (35 °C). The boundary conditions for the discrete phase were set to let the droplet escape when inhaled from the noses or extracted for the HVAC grilles, whereas they are trapped (i.e. eliminated from the domain) when touching a manikin or a surface.

**Figure 3:**
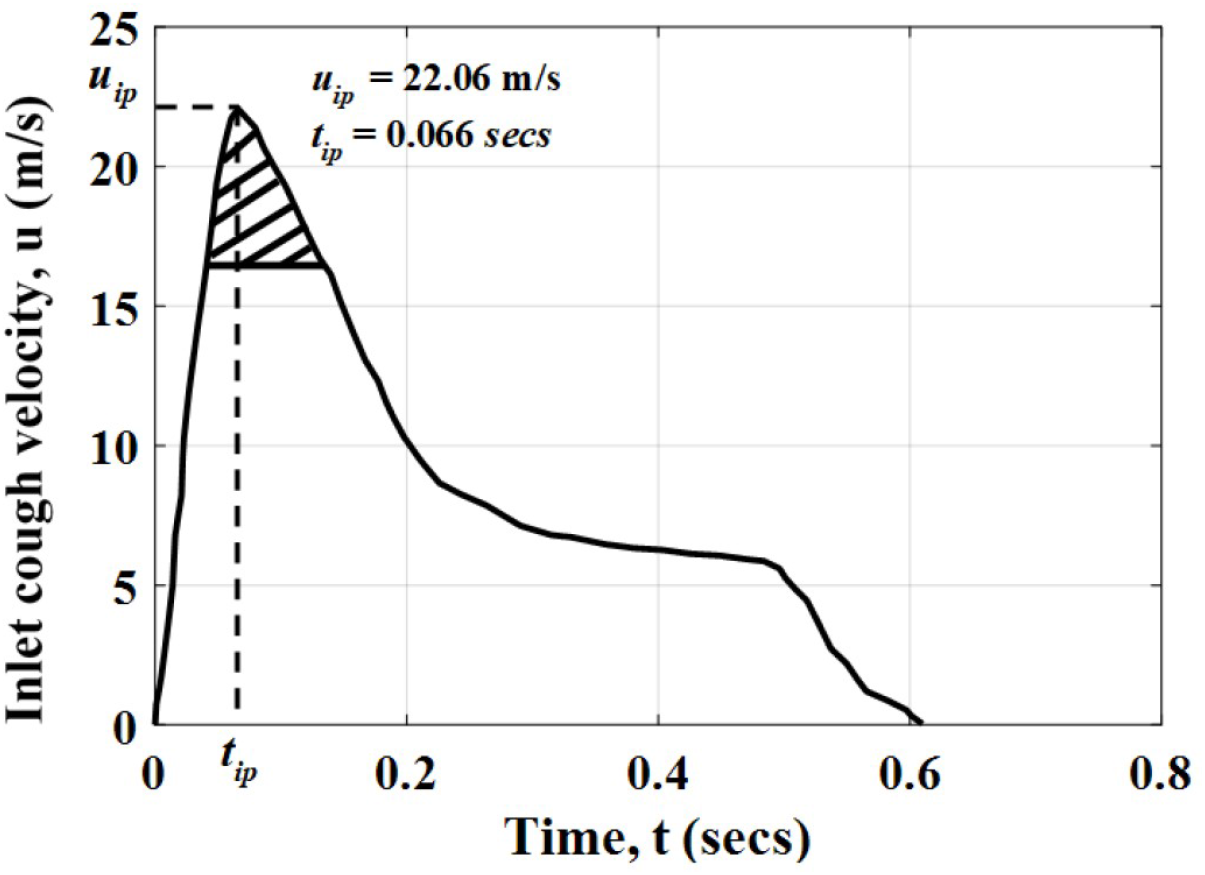
Temporal evolution of the inlet cough velocity (eulerian phase) and droplet injection (lagrangian phase, shaded part) (reproduced from Gupta et al. (2009)).

For what concerns the Eulerian phase, most of the studies in the literature deals with RANS turbulence models to account for the turbulence effects on the flow and droplets, in both a steady (RANS) or unsteady (URANS) fashion. The exploitation of a direct solution of the turbulent flow, or at least of the largest scales, (as done in LES) showed some promising improvements (see for example [22]), but it is still not widespread due to computational effort involved. Also in this work, the URANS approach was chosen due to the complexity of the cases investigated. The turbulence effects were modeled with a k-ε RNG model (as already done in previous works [19], [25]–[27]) and scalable wall functions.

The inhalation/exhalation of the “target” manikin was modeled with pressure-outlet boundary conditions with a target mass flow rate. The literature contains much information, even though scattered, about the transient respiration process, usually described as a sinusoidal function [19]-[21].

However, the approach proposed by Gao et al. [25] was followed, who modeled it as steady inhalation with a flow rate of 0.14 l/s. The overall opening area of the nose is 0.56 cm^2^. For the sake of simplicity, such flow rate was kept equal for both men and kids.

To track the dispersion of the contaminated fluid, a passive transported scalar was added to the simulations, with a mass diffusivity ρD equal to 2.5e-5 kg/m s. In terms of boundary conditions, a value of 1 was applied to the mouth of the spreader subject, whereas a zero-flux condition was applied to all the outlets.

In an attempt to provide a more quantitative characterization of the results in the different scenarios, the approach proposed by Gao et al. [25] was followed. They described the fraction of the sneezed air in the inhaled air using the Infection Index η, described by the equation:

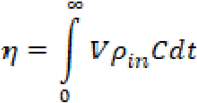

Where *V* is the inhalation rate [m^3^/s], *ρ_in_* is the inhalated air density [kg/m^3^] and C the mass fraction of the coughed air in the inhaled air. The parameter provides a representation of the integral amount of contaminated air inhaled by the subjects during the temporal evolution of the event.

The computational grid was generated in ANSYS Meshing and consisted of a tetrahedral mesh with a 5 prismatic layers on the walls, which is adequate given the limited role played by viscous effects on the flow. Then the mesh was converted in polyhedra in ANSYS Fluent, resulting in 3.87 million polyhedrons. The mesh is depicted in Figure 4, where only the Case 1 is reported for the sake of brevity.

**Figure 4:**
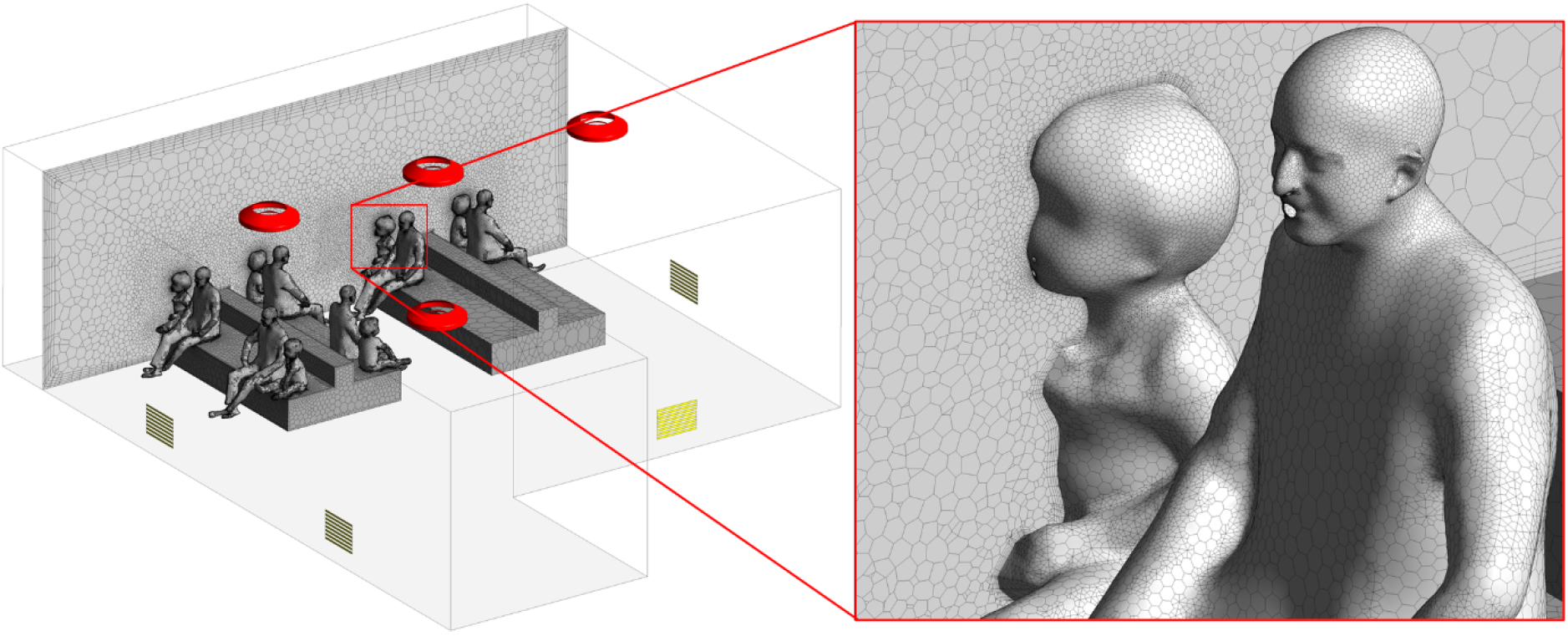
Details of the computational grid used in the simulations of Case 1

The solution was obtained with a pressure-based solver and a SIMPLE algorithm with a Second Order scheme for pressure, Second Order Upwind for all variables (except for turbulence quantities), while time was treated with a Bounded Second Order Implicit scheme. A constant time step of 10^-2^ s was chosen, with 10 sub-iterations, which are appropriate to achieve a sufficient reduction in the residuals within the time step. The process was solved with a transient simulation of the overall duration of 60 seconds for both the waiting room (Case 1) and the recovery room (Case 2).

## Results

### Case 1 (Waiting Room)

The quantitative results of the simulations for the waiting room are plotted in Figure 5. As already mentioned, they consist in scenario A (without HVAC), scenario B (HVAC with nominal air-flow of 2020 m^3^/h for each diffuser) and scenario C (HVAC with a doubled airflow of 4040 m^3^/h). The x-y plots represent the temporal evolution of the index η, which describes the cumulative amount of contaminated air inhaled by different subjects as a consequence of the coughing event. Units are micrograms (μg). Such comparison allows studying the airborne contagion exposure for all the people sitting in the room. In this work we focused particularly on children’s contagion exposure. The infection index is particularly influenced by the HVAC airflow as well as the subject positions in the room relatively to the spreader’s position.

**Figure 5:**
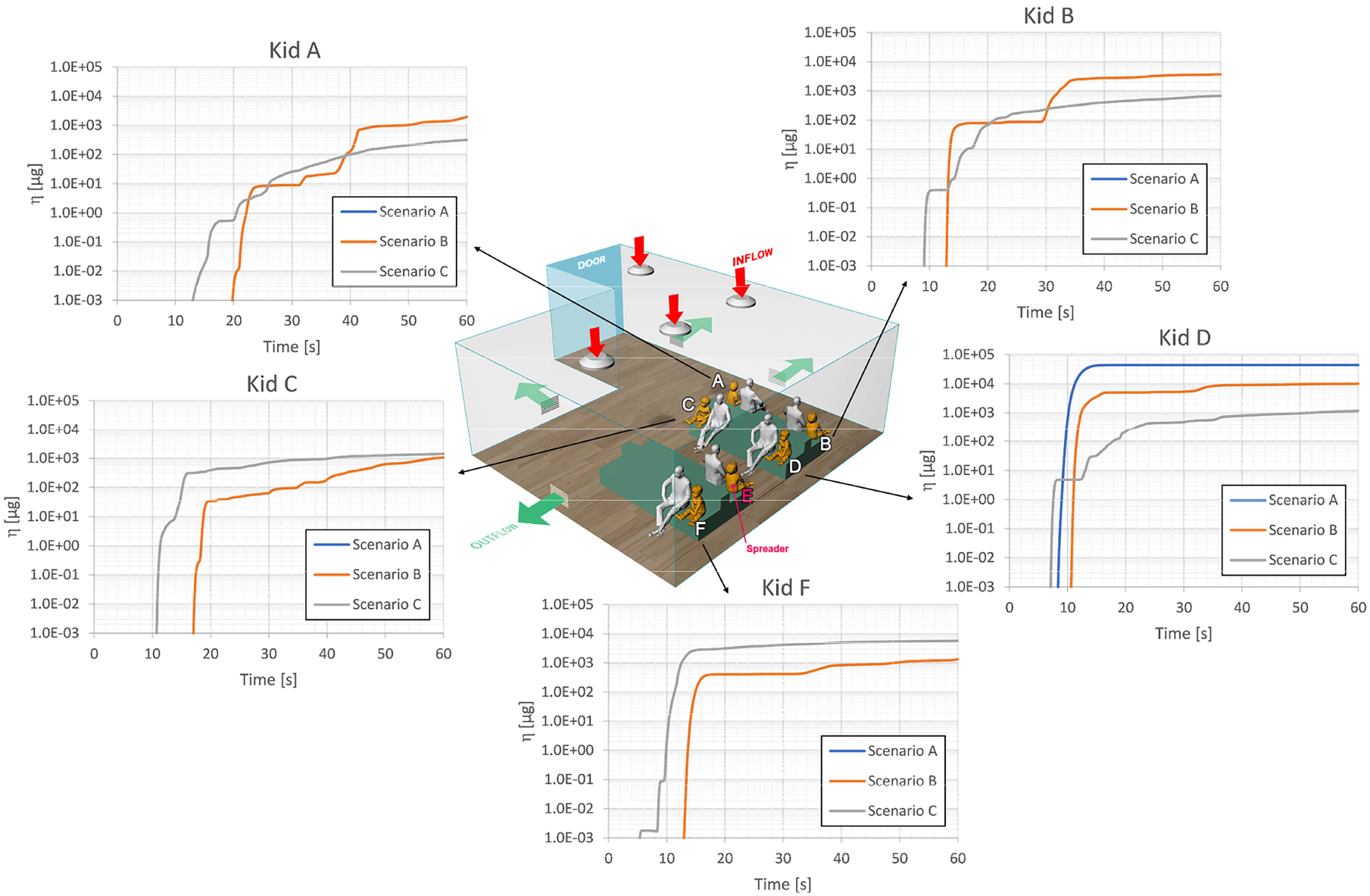
Temporal evolution of the Infection Index η for the kids in the waiting room.

Looking at the charts it is possible to notice that in absence of HVAC airflow the contaminated air coughed by the spreader subject reaches only the subject in front (Kid D), at least in the time interval considered (60 s). When the HVAC system is considered the stronger the intensity of the airflow, the lower the concentration that affects the Kid D with a percentage reduction of -99.6% of concentration in Scenario C respect to Scenario A. Nonetheless, the increase in HVAC flow rate (thus in flow velocity) leads to a greater dispersion of the contaminated air far from the spreader subject. It is the Scenario C in which η reaches relevant values before the other. This is indicative of a more intense turbulent transport throughout the room.

With the aim of better understanding from a qualitative point of view the results just described, images at t=1 s and 5 s were selected to show the impact of the HVAC operating conditions in the very firsts seconds after the start of the coughing event. Looking Figure 6, several considerations can be raised. As it is possible to observe, droplets beyond 100 μm are subject to gravitational force and drop within 1 m irrespective of the intensity of the HVAC airflow. Only in the Scenario C the flow from the diffusers is strong enough to let them be pulled back to the face of the spreader subject. Droplets with a smaller diameter are insensitive to gravity and are transported by the coughed stream of contaminated air.

**Figure 6:**
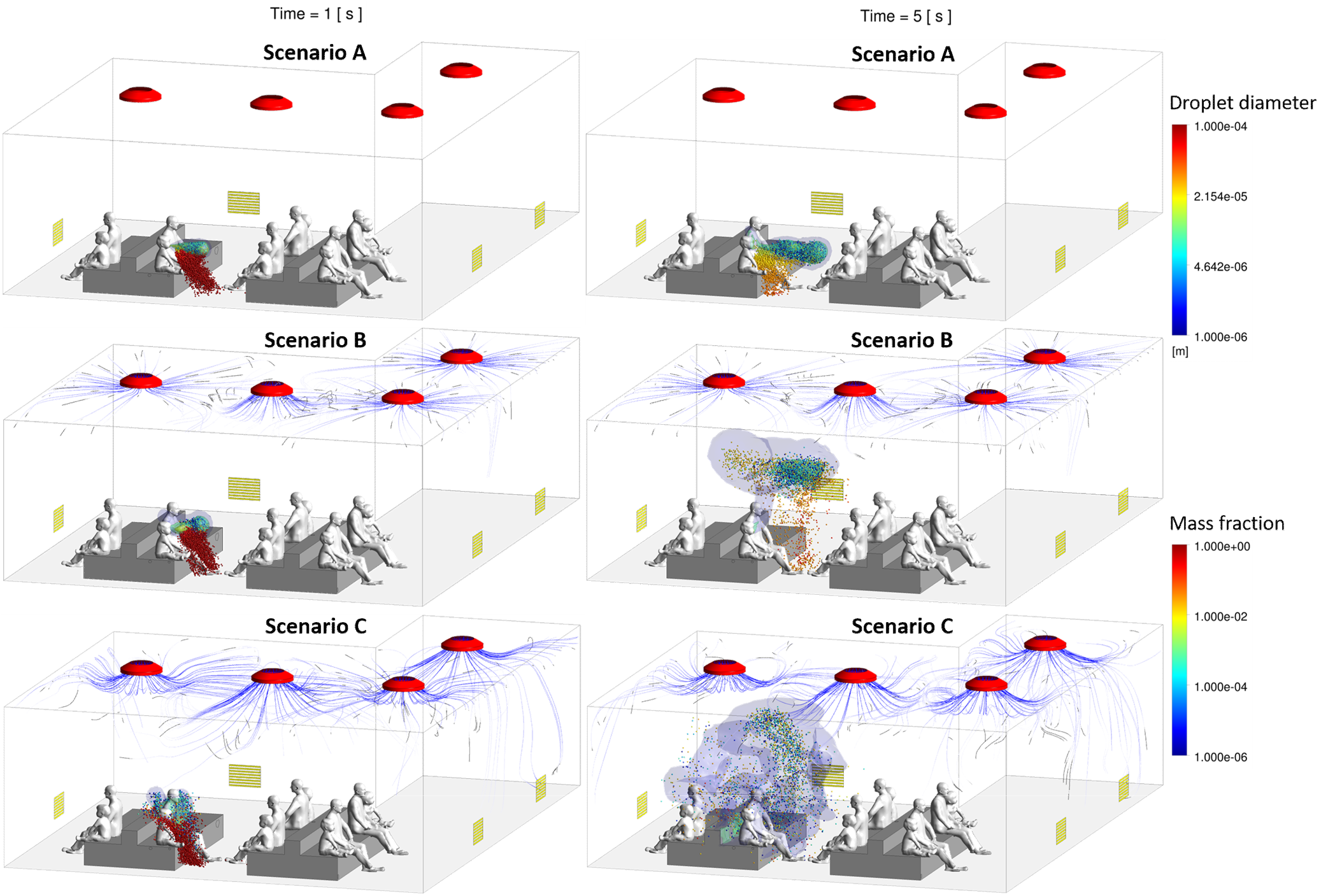
Prospective view of the Scenari A, B and C at t=1 s (left) and 5 s (right). The spheres represent the droplets colored by the diameter size (top right legend). The contaminated air is represented by different iso-surfaces colored by mass fraction.

The development over time of the situation is visible at t=5 s, for which in the Scenario A heaviest droplets are progressively dragged to the ground while the lightest droplets (diameter approximately lower than 10 μm) continue their travel unhindered until they will reach the subject in front (Kid D). In presence of an HVAC (Scenario B), as anticipated, the coughed stream loses rapidly its momentum and is captured by a recirculation zone created by the HVAC between the spreader subject and the lateral wall (see supplementary materials: “Case 1 Video” and “Contaminant Recirculation Video”). Interestingly, for the Scenario C the further increase in the HVAC airflow promotes the turbulent transport, resulting in a complete disruption of the droplet cloud and an enhancement of its dispersion within the room.

To provide additional evidence of the impact of the HVAC airflow on the droplet dynamics, the temporal evolution of their total mass in the room was calculated and plotted in Figure 7. All the curves start from the same condition of 0.72 g, as prescribed in the setup of the discrete phase injection. Then, the mass suddenly drops by 2 orders of magnitude in the 5 s following the start of the event. Such a significant reduction is associated to the droplets of large diameter (and thus large mass), which are more influenced by the gravitational force. Interestingly, negligible differences are present between Scenario A and B, confirming that the HVAC has a minimal effect in the first phase of the event. On the contrary, the Scenario C returns a more significant reduction in mass. This effect may be ascribed to the HVAC airflow that pushes the droplets back to the spreader subject, where they are trapped on its surface. In the second phase of the simulation (5 s ≤ t ≤ 60 s) the evolution shows a significant slowing-down, since after the impact with the subject in front (Kid D) the droplets continue to fall to the ground and to be transported either by buoyancy (Scenario A) or turbulent transport (Scenario B and C). It is however worth highlighting how at the end of the time under investigation Scenario B is characterized by a greater amout of droplet mass respect to Scenario A and Scenario C. This result is achieved because, while in Scenario A most of the haviest droplets fall to the ground, in Scenario B they follow the HVAC air flow for the entire time considered. In Scenario C droplets are more diluited and their concentration decreaseas more than in the other two scenarios.

**Figure 7:**
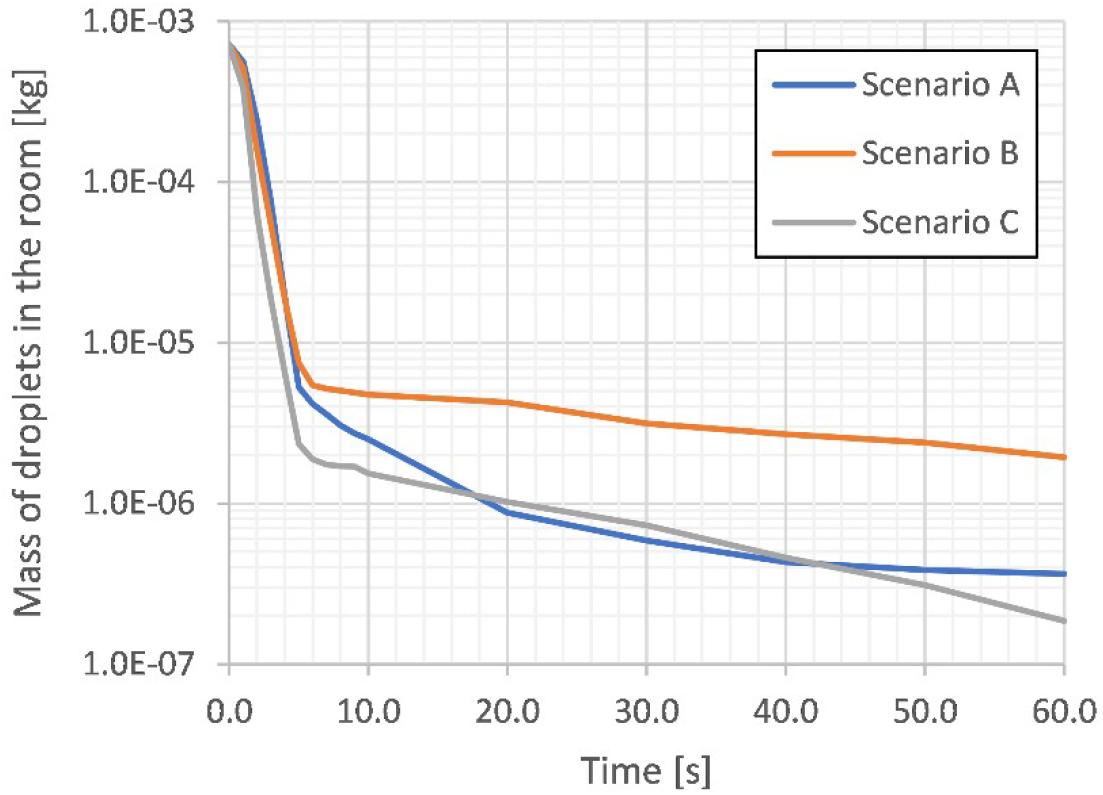
Temporal evolution of the mass of the droplets in the waiting room.

It is reasonable to think that in absence of HVAC the smallest droplets survive only thanks to buoyancy, whereas the largest one are progressively dragged to the ground. On the contrary, the reduction rate in presence of HVAC is reduced by the turbulent transport throughout the room, which is far more intense than buoyancy and gravitational effects, preventing also the large droplets to fall down.

To better understand the conditions at the middle of the simulations (t=30 s), it is possible to refer to Figure 8. For the Scenario A, about an half of the people sitting in the room are not reached by the droplet cloud and two columns of droplets are formed above Kid E and D as a consequence of the temperature-driven buoyancy, which is associated to very low-speed motion.

**Figure 8:**
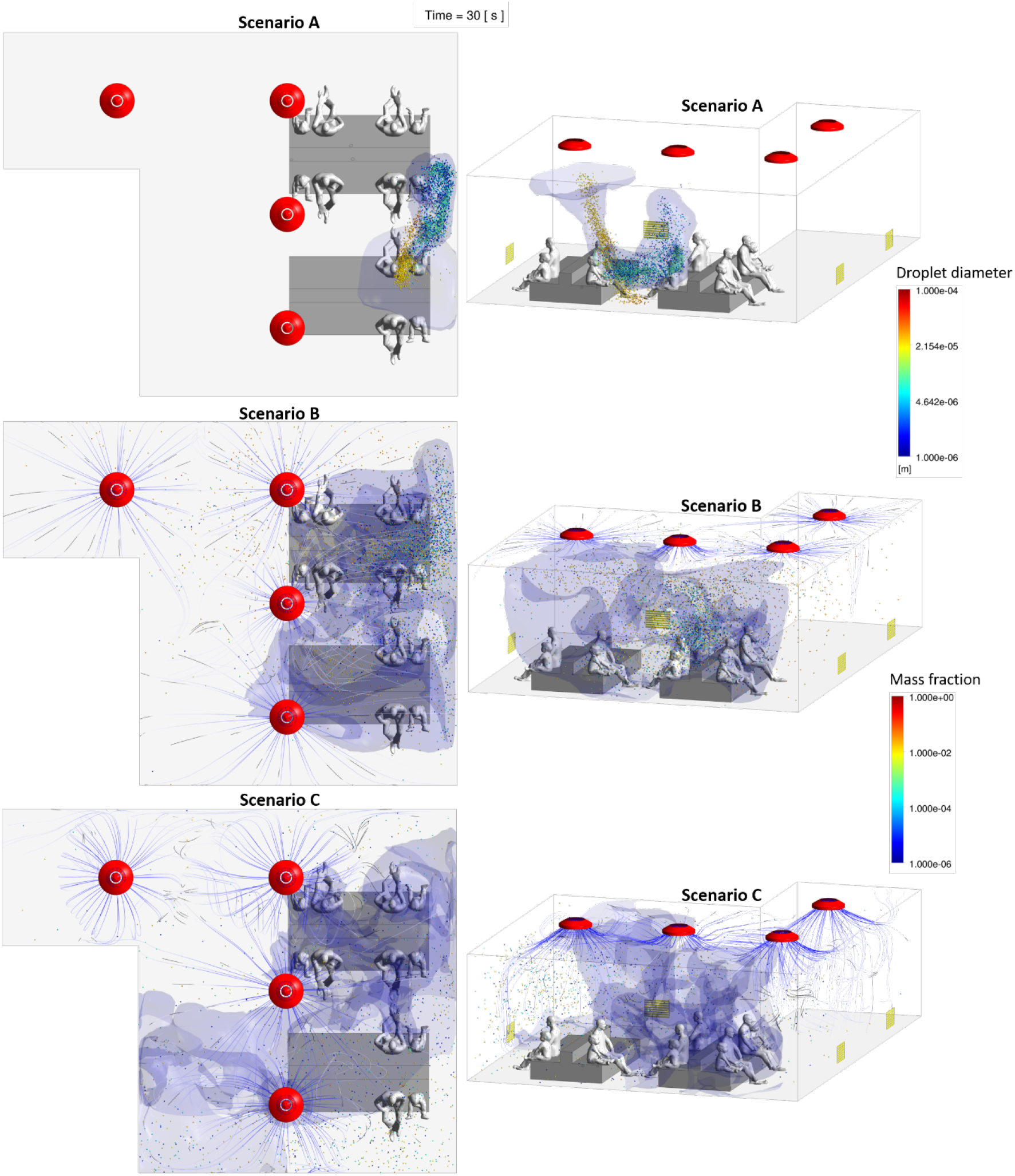
Top (left) and prospective (right) views of the Scenari 0, 1 and 2 at t=30 s. The spheres represent the droplets colored by the diameter size (top right legend). The contaminated air is represented by different iso-surfaces colored by mass fraction (bottom right legend). The streamlines originating from the HVAC inlet diffusors show the air velocity.

In the Scenario B and C, where the HVAC system is activated, the cloud of droplets covers considerable distances inside the room and reaches all the people inside. In these two scenarios, all the people in the room come into contact with the dispersed airborne contaminant and the droplets cloud assumes a turbulent motion relatively of the HVAC airflow settings. As highlighted by the figures, the droplet spreading is more intense as the HVAC flow rate increases.

### Case 2 (Recovery Room)

In Case 2 a time interval of 60 s was simulated to allow the contaminated air to reach the other subject in the room (Kid A). Unlike the waiting room (Case 1), the propagation speed of both contaminated air and droplets is significantly reduced due to the presence of LEV system and for different trajectory of the coughing event, which is directed towards the ceiling. It is worth underlining that, contrary to Case 1, HVAC is present in both scenarios, but with a smaller flow rate entering from the diffuser (approximately a quarter of the previous nominal condition). In Case 2 we studied two different conditions: Scenario 0, where only the HVAC system is present, and Scenario 1, which also considers a LEV system positioned directly above the face of the spreader subject (Kid A).

By analyzing the simulations, it is possible to appreciate immediately the advantage of equipping the recovery room with a LEV system. A more quantitative comparison is reported in Figure 9, which describes the temporal evolution of the Infection index. Considering that the same range of Case 1 was used, it is possible to notice that the amount of contaminated air inhaled is significantly lower than what observed in the waiting room. As already mentioned, this can be ascribed to the different trajectory of the coughed stream as well as the lower HVAC flow rate. More importantly, the LEV system provides a virtually complete abatement of contamination.

**Figure 9:**
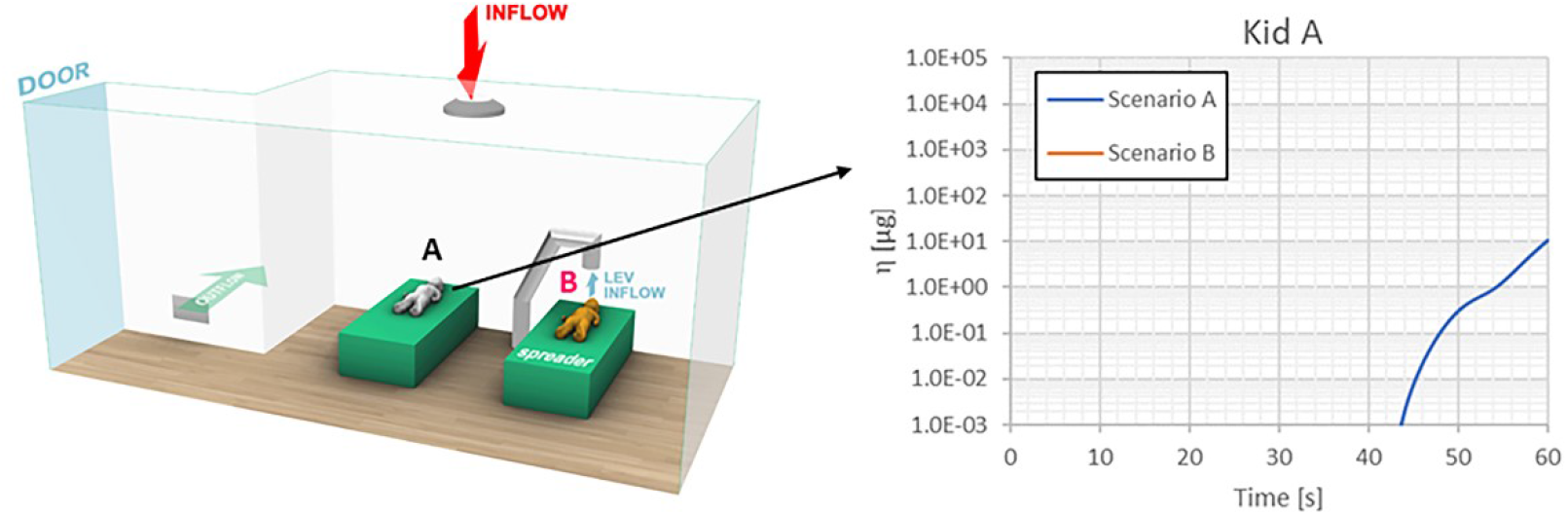
Temporal evolution of the Infection Index η for the kids in the recovery room.

A more qualitative description of the results of the coughing event is reported in Figure 10 where the two scenarios are compared similarly to what already done for the Case 1. In the absence of any LEV system (Scenario A), the contaminated air and the droplets are transported at first by a combination of cough and temperature-driven buoyancy. Subsequently, the HVAC airflow dominates and enhances the turbulent transport throughout the room. Considering instead the Scenario B, where the LEV system is present, coughed air and droplets are immediately captured and extracted from the room, thus hindering any further spreading and contamination of other subjects (see Supplementary Materials: Case 2 Video).

**Figure 10:**
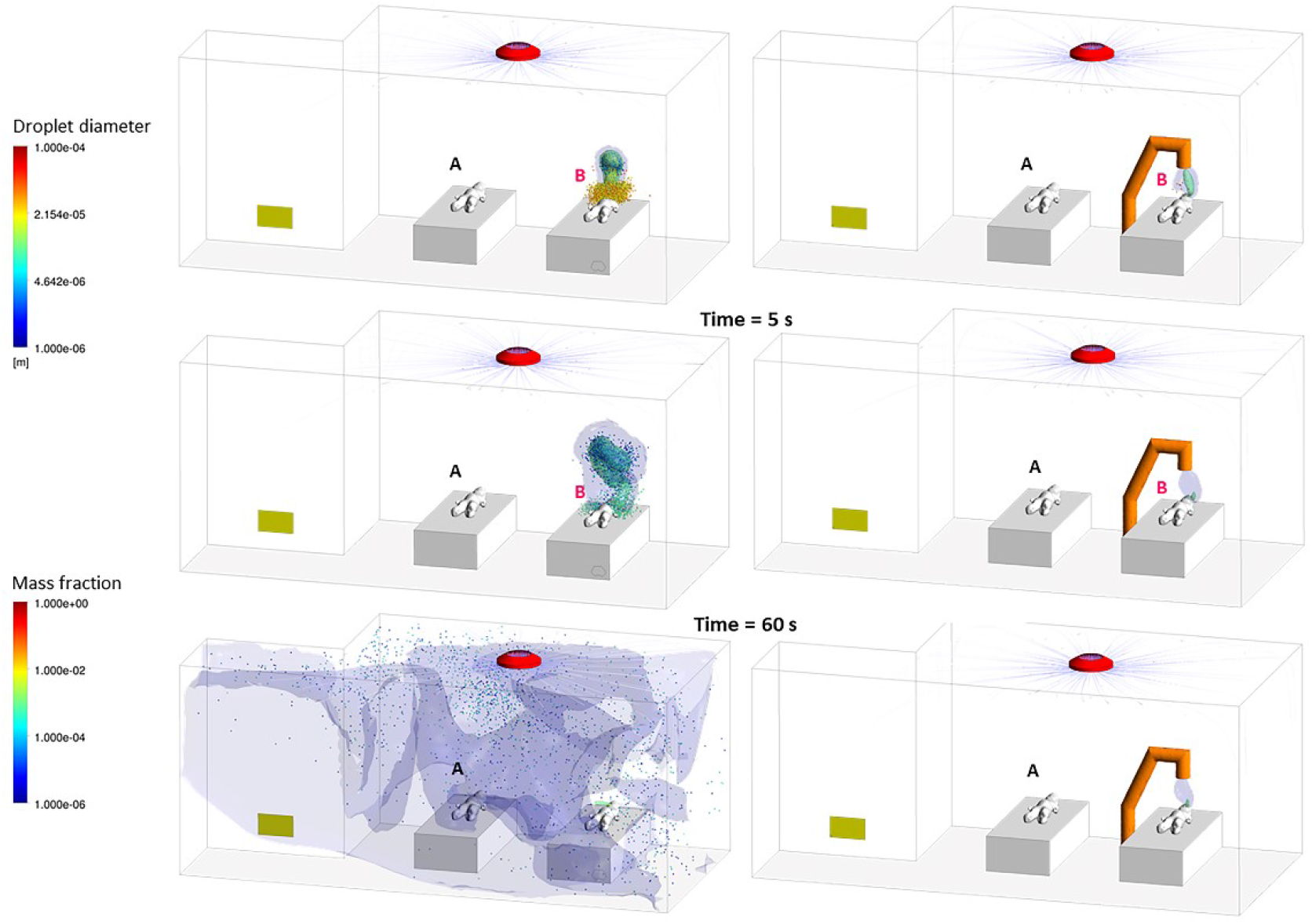
Top and prospective view of the recovery room for the Scenario 0, 1 and 2 at t=60 s. The spheres represent the droplets colored by the diameter size (top right legend). The contaminated air is represented by different iso-surfaces colored by mass fraction (bottom right legend). The streamlines originating from the HVAC inlet diffusors show the air velocity.

The effectiveness of the LEV system is confirmed also by the temporal evolution of the droplet mass in the room (reported in Figure 11). As already showed, in the Scenario B almost all the droplets are extracted within 2 s, while in the Scenario A are free to propagate throughout the room. Unfortunately the analysis of contagion risk from the airborne contaminants within hospital’s rooms through the CFD approach is no generalizable as it is dependent on numerous specific factors such as the room layout, the HVAC diffusors and return grilles position, the people position in the room and their dynamic physical activities. However the CFD is a very useful tool for assessing and predicting the viral infection risk within closed places by running a customized CFD project based on each specific room.

**Figure 11:**
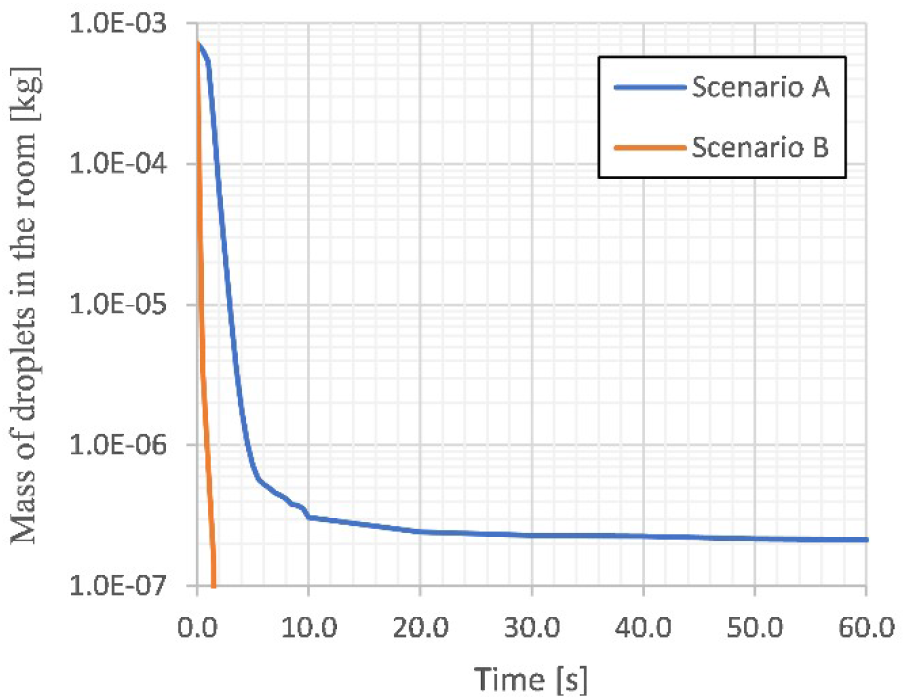
Temporal evolution of the mass of the droplets in the recovery room.

## Conclusions

Our study shows us that the HVAC system in a closed waiting room generates a marked space diffusion of droplets from a chough event. Although the doubling of HVAC’s air flow allows for a marked reduction in the airborn contaminant concentration this also leads to a substantial increase of turbulent air motions resulting in an increased and faster long-range spreading of droplets and air contaminants in the room. In indoor environment, the doubling of the air conditioned flow allows a relevant (up to 77%) reduction in droplets mass concentration within the room compared to nominal air flow. In the first 5 seconds from the cough event, no particular differences in droplets concentration are detectable in the room, while the relevant differences occur within 25 seconds due to the air conditioned systems that induce droplets air dispersion. In absence of air conditioning, larger particles more easily settle on the ground, while for nominal or doulbe-flow HVAC air-flow even the largest particles are subjected to turbulent phenomena and follow the air conditioned flow remaining suspended for longer time. Despite that the calculation of Infection Index for all the subjects in the room proved that a higher air exchange allows to reduce in the long term the overall amount of inhaled contaminated air. Further studies are needed to deepen the contribution of natural ventilation to the scenarios analysed to understand if this can contribute to contaminants exposure reduction for the people inside the rooms.

In the simulations performed in recovery rooms, the first 40 seconds from the cough event are not sufficient to allow both droplets and contaminated air to reach the Kid A. After that, in the absence of a Local Exhaust Ventilation system, the HVAC air-flow favours their turbulent dispersion within the room. The presence of a LEV unit placed above the patient’s face shows a very high capacity to reduce droplets and contaminated air in the room, ensuring a total absence of exposure to contagion risk for the patient A. In our simulations based on quantitative CFD data, LEV system is able to remove both droplets and contaminated air in the first seconds immediately following the cough event. The development of CFD 3D simulations in healthcare environments can provide a qualitative and quantitative prediction of contagion risk and valuable help in optimizing the prevention processes aimed at minimizing infectious risk for patients and healthcare professionals during a pandemic.

## Data Availability

The authors confirm that the data supporting the findings of this study are available within the article and its supplementary materials.

## Aknowledgments

Authors are grateful to the RESCOP study group established to analyse the possible link between indoor/outdoor air pollution and COVID-19 outbreaks. The Italian Society of Environmental Medicine (SIMA) is grateful to Falck Renewables for its support.

## Authors Contribution

Luca Borro, Lorenzo Mazzei, Massimiliano Raponi, Prisco Piscitelli, Alessandro Miani, Aurelio Secinaro conceived and designed the study;

Luca Borro, Lorenzo Mazzei, and Aurelio Secinaro performed the simulations.

All the authors contributed to write and revise the manuscript.

## Funding Information

No external funding received

## Disclosure Statement

All authors declare no conflict of interest

Patient and Public Involvement Statement:

No patients were involved in this study

## Nomenclature

C: Mass fraction of contaminated air [-]

D: Molecular diffusivity [m^2^/s]

k: Turbulence kinetic energy [m^2^/s^2^]

t: Time [s]

T: Temperature [C]

V: Volume rate [m^3^/s]

Acronyms

CFD: Computational Fluid Dynamics

HVAC: Heating, Ventilation and Air Conditioning

LEV: Local Exhaust Ventilation

MFR: mass flow rate

RANS: Reynolds-Averaged Navier-Stokes

URANS: Unsteady RANS

Greeks

ε: Turbulence dissipation rate [m^2^/s^3^]

η: Infection index [μg]

ρ_in_: Density of inhaled air [kg/m3]

## Notes

### Competing Interest Statement

The authors have declared no competing interest.

### Author Declarations

Bambino Gesu Childrens Hospital - Scientific Directorate

